# Seroprevalence screening of chronic *Aspergillus* infection in a post-tuberculosis cohort in Senegal: A cross-sectional study comparing ELISA and rapid diagnostic tests

**DOI:** 10.1101/2025.11.19.25340617

**Authors:** Touré Mariama, Ndiaye Pape Ibrahima, Sow Djiby, Diallo Mamadou Alpha, Diongue Khadim, Seck Mame Cheikh, Ndiaye Mouhamadou, Ndiaye Daouda, Ndiaye Faly DIOP, Fortes Louise, Denning David, Badiane Aida Sadikh

## Abstract

**Background:** Chronic Pulmonary Aspergillosis (CPA) is a significant, yet often overlooked, sequela of pulmonary tuberculosis (TB), particularly in resource-limited settings. Data on the seroprevalence of *Aspergillus* infection in Senegal is absent, and diagnostic capacity is limited. This study aimed to determine the seroprevalence of *Aspergillus*-specific antibodies among patients with a history of TB in Senegal and to evaluate the performance of a Rapid Diagnostic Test (RDT) against an Enzyme-Linked Immunosorbent Assay (ELISA).

**Methods:** A cross-sectional study was conducted at two health centers in Senegal; Wakhinane and Yeumbeul among patients with chronic respiratory symptoms in Senegal. Each participant provided a blood sample tested for *Aspergillus* antibodies using both an RDT and an ELISA. Results were classified as positive, negative, invalid, or not tested. Prevalence and agreement between the two assays were calculated using valid results only. Demographic data were collected, and descriptive statistics with test performance analyses were conducted.

**Results:** The overall seroprevalence was significantly higher by ELISA (11.9%; 38/320) than by RDT (5.5%; 11/200). Site-specific analysis revealed disparities: Wakhinane showed higher RDT positivity (4.5% vs. 1.0%), while Yeumbeul had higher ELISA positivity (13.5% vs. 11.2%). The cohort was predominantly male (66.0%) with a median age of 30 years. ROC analysis of the ELISA identified an optimal threshold that balanced sensitivity (78%) and specificity (89%).

**Conclusion:** This study provides the first serological evidence of substantial *Aspergillus* antibody prevalence among post-TB patients in Senegal, suggesting a significant burden of undiagnosed CPA. The higher sensitivity of ELISA makes it essential for surveillance and confirmation, while the RDT’s practicality offers a viable option for initial screening in peripheral clinics. These findings underscore the urgent need to integrate CPA diagnosis into routine post-TB care and to strengthen fungal diagnostic capacity in West Africa.

## Introduction

Chronic pulmonary aspergillosis (CPA) is a serious and underdiagnosed fungal disease caused mainly by *Aspergillus fumigatus*. The major clinical forms differ in terms of the level of immunosuppression and underlying pulmonary lesions [1;2]. It typically develops in individuals whose lungs have been damaged by previous infections such as tuberculosis (TB). Based on estimation, the CPA has an incidence of 1 390 766 for new CPA cases from newly presenting tuberculosis cases (with 153 029 deaths, 12·5% of overall tuberculosis deaths). In addition, 446 505 new CPA cases are estimated to have presented in 2020 in people who had pulmonary tuberculosis in the previous 5 years, an incidence of 1 837 272 new cases (23·6 per 100 000 population). The associated mortality rate ranges from 20% to 7% depending on comorbidities and access to antifungal drugs [3;4].

Behind the global struggle against tuberculosis lies a hidden and devastating aftermath. For millions of people who have survived TB, their lungs can be left scarred and vulnerable, creating a fertile ground for a relentless fungal infection known as CPA. The scale of this silent crisis is staggering. A 2024 analysis estimates that nearly 1.4 million new CPA cases arise annually from recently diagnosed tuberculosis patients, leading to approximately 153,000 deaths accounting for 12.5% of all TB mortality [5]. The lethality of this condition is further underscored by a subsequent 2024 report, which confirms the profoundly high mortality of CPA, with case-fatality rates reaching up to 50% within five years, solidifying its status as a major, yet often unrecorded, cause of post-TB mortality [6]. Furthermore, an additional 447,000 new CPA cases emerge each year among individuals who had pulmonary TB within the preceding five years, culminating in a total incidence of 1.84 million new cases globally [5]. This is not a minor footnote to TB; it is a lethal sequela demanding urgent integration into mainstream respiratory health initiatives.

Recognizing CPA is challenging, especially in countries where TB is widespread. Both conditions cause chronic cough, fatigue, and lung damage, leading to frequent misdiagnosis. In such cases, detecting *Aspergillus*-specific antibodies plays a crucial role in confirming CPA [2].

ELISA tests are widely used for this purpose and have proven reliable [7]. However, they require laboratory infrastructure, electricity, and trained staff resources that are often scarce in African hospitals. Rapid diagnostic tests (RDT), such as lateral flow assays, have emerged as a more practical option. They can be performed on-site, provide results in minutes, and are particularly suited for low-resource settings [8; 9].

Despite their potential, data on RDT performance in Africa remain limited. In Senegal, where TB is highly endemic, no published data exist on the prevalence of *Aspergillus*-specific antibodies among patients suspected of CPA. Generating such evidence is essential to improve diagnosis, clinical awareness, and policy development for fungal infections.

The goal of this study was to estimate the prevalence of *Aspergillus* antibody positivity using RDT and ELISA in Senegalese patients with chronic respiratory symptoms.

## Methods

### Study design and participants

A cross-sectional study was conducted among adult patients diagnosed with pulmonary tuberculosis who had completed the fifth or sixth month of anti-tuberculosis treatment. Recruitment took place at two primary healthcare centers located in the suburban districts of Dakar, Senegal; Wakhinane and Yeumbeul areas known for a high burden of tuberculosis. Eligible participants were those nearing the end of their TB treatment course and attending routine follow-up visits. Written informed consent was obtained from all participants prior to inclusion in the study.

### Laboratory procedures

Serum, isolated from centrifuged blood samples collected from each patient, was stored at - 20°C until subsequent analysis with two serological assays. For each patient a RDT which is a lateral flow assay designed to detect *Aspergillus*-specific IgG/IgM antibodies (LDBio) were performed.

For the rapid detection of *Aspergillus*-specific antibodies, the Aspergillus ICT IgG-IgM lateral flow assay (LD BIO, Lyon, France) was used. This immunochromatographic test was performed according to the manufacturer’s instructions. Briefly, we applied serum samples to the test device followed by the provided buffer solution. The results were interpreted after 10 minutes of incubation at room temperature. The presence of both control and test lines indicated a positive result for *Aspergillus* antibodies, while the appearance of only the control line indicated a negative result.

Serum samples were analyzed for *Aspergillus*-specific IgG antibodies using a commercial ELISA kit (Abcam, ab247190) at the International Research & Training Center on Genomics and Health Surveillance (CIGASS). The results were assessed independently by two technicians.The optical density was measured at 450 nm. For each sample, the average absorbance was calculated and compared to the average absorbance of the kit’s cut-off control. This ratio was then converted into arbitrary Units (U) using the formula: Units (U) = (Average OD of Sample / Average OD of Cut-off Control) × 10. Following the kit’s guidelines, an antibody concentration of ≥11 U/mL was considered as a positive result, indicating a specific serological response to *Aspergillus*. The result was equivocal when the antibody concentration was between 9 and 11 U/ml and negative when < 11 U/ml. Only valid results were included in the final analysis.

### Data management and statistical analysis

Data were entered into a structured spreadsheet in Excel (Version 16.102.2) and analyzed using R (version 4.5.1). Prevalence was calculated as the proportion of positive results among patients tested with each assay (n = 201 for RDT; n = 320 for ELISA). Invalid and untested results were excluded from denominators.

For patients who had valid results for both tests, cross-tabulations were calculated to assess agreement. Cohen’s kappa coefficient was used to quantify concordance between RDT and ELISA.

### Ethical considerations

The study protocol was approved by the Ethic committee (CNERS) of the ministry of health (SEN15/46). All participants provided written informed consent prior to enrolment.

## Results

### Study Population

A total of 320 participants were recruited from the two health centers of Wakhinane (n=224) and Yeumbeul (n=96). The overall study population was relatively young, with a mean age of 32.5 years (±12.6 SD) and a median age of 30.0 years (IQR 23.0-40.0). The population from Yeumbeul was slightly younger (median 27.5 years) compared to Wakhinane (median 30.0 years). Males constituted the majority of the cohort (66.0%, 198/320), with a significantly higher proportion of male participants in Yeumbeul (75.3%) compared to Wakhinane (62.8%).

It is important to note the presence of missing data, particularly for age (n=77) and sex (n=20), with a higher volume of missing sex data from the Yeumbeul site (n=19). These gaps were accounted for in the denominator of the respective analyses (Table 1).

**Table 1:**
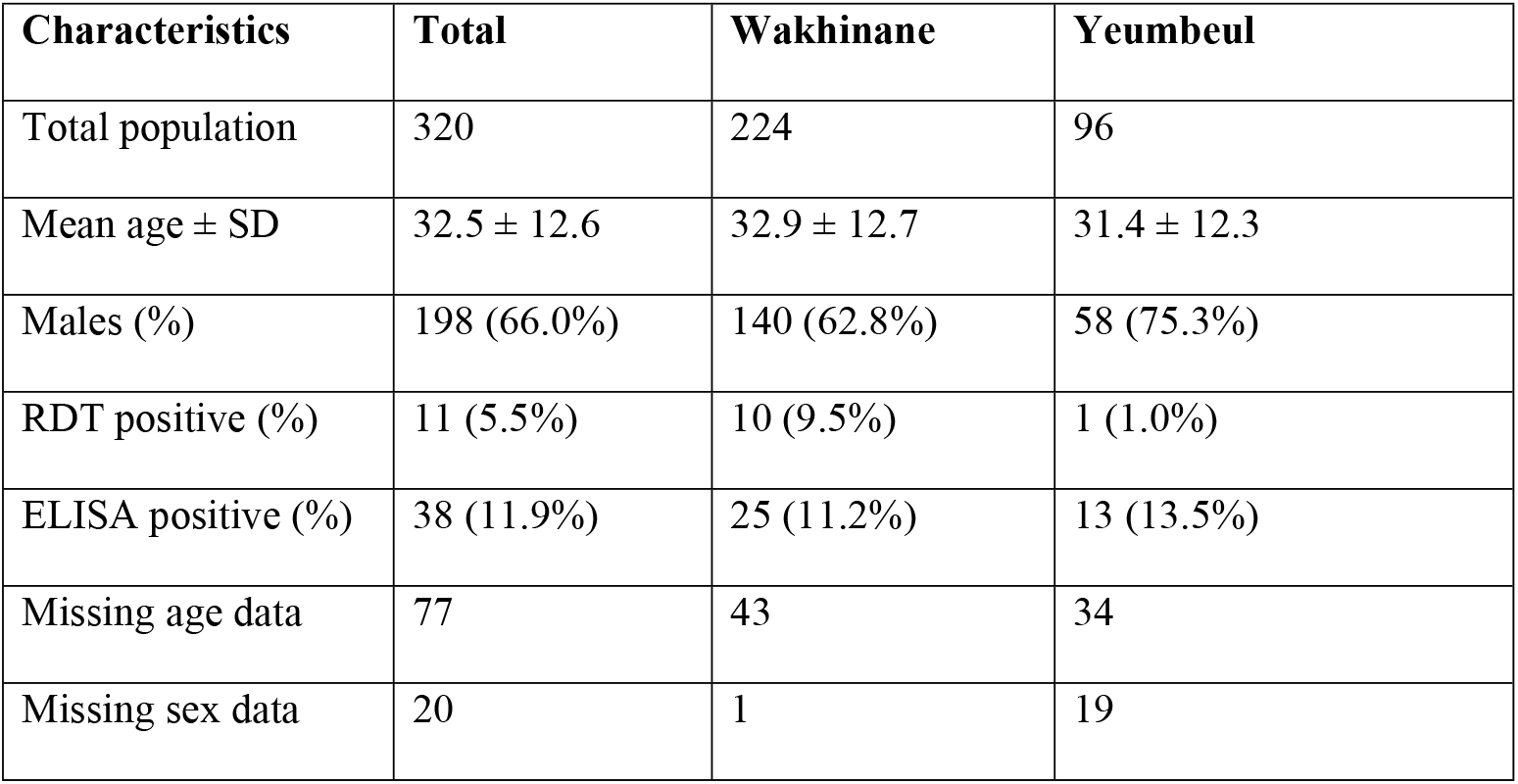
Characteristics of the study population and the tests performed.

| Characteristics | Total | Wakhinane | Yeumbeul |
| --- | --- | --- | --- |
| Total population | 320 | 224 | 96 |
| Mean age ± SD | 32.5 ± 12.6 | 32.9 ± 12.7 | 31.4 ± 12.3 |
| Males (%) | 198 (66.0%) | 140 (62.8%) | 58 (75.3%) |
| RDT positive (%) | 11 (5.5%) | 10 (9.5%) | 1 (1.0%) |
| ELISA positive (%) | 38 (11.9%) | 25 (11.2%) | 13 (13.5%) |
| Missing age data | 77 | 43 | 34 |
| Missing sex data | 20 | 1 | 19 |

### Serological results

Among the 320 enrolled patients, all were successfully tested for fungal markers using ELISA. Rapid Diagnostic Test (RDT) results were available for a subgroup of 200 participants. The unavailability of RDT results for the remaining 118 individuals was primarily due to a limited supply of test kits, which were exhausted during the recruitment period.

The seroprevalence of fungal antigens or antibodies, as detected by different diagnostic methods, varied between the sites. Using a Rapid Diagnostic Test (RDT), the overall positivity was 5.5% (11/200). A marked disparity was observed between the health centers: Wakhinane had an RDT positivity rate of 9.5% (10/105), which was substantially higher than the 1.0% (1/95) observed in Yeumbeul (Table 1).

Conversely, the more sensitive ELISA test revealed a higher overall seroprevalence of 11.9% (38/320). The site-specific analysis using ELISA showed a different pattern from the RDT results. The seroprevalence in Yeumbeul was 13.5% (13/96), which was slightly higher than the 11.2% (25/223) found in Wakhinane.

Seropositivity was defined by both tests positive and/or ELISA positive according to the manufacturer’s recommended cutoff.

### Agreement Between RDT and ELISA

Among the 201 individuals who underwent both tests, discordant results were observed in approximately 28.9% of paired samples, mainly in patients who tested negative by RDT but positive or equivocal by ELISA.

A receiver operating characteristic (ROC) analysis was performed to determine the optimal optical density threshold for the ELISA test, balancing sensitivity and specificity against the reference standard. The relationship between the ELISA threshold and key diagnostic metrics is detailed in Figure 1.

**Figure 1:**
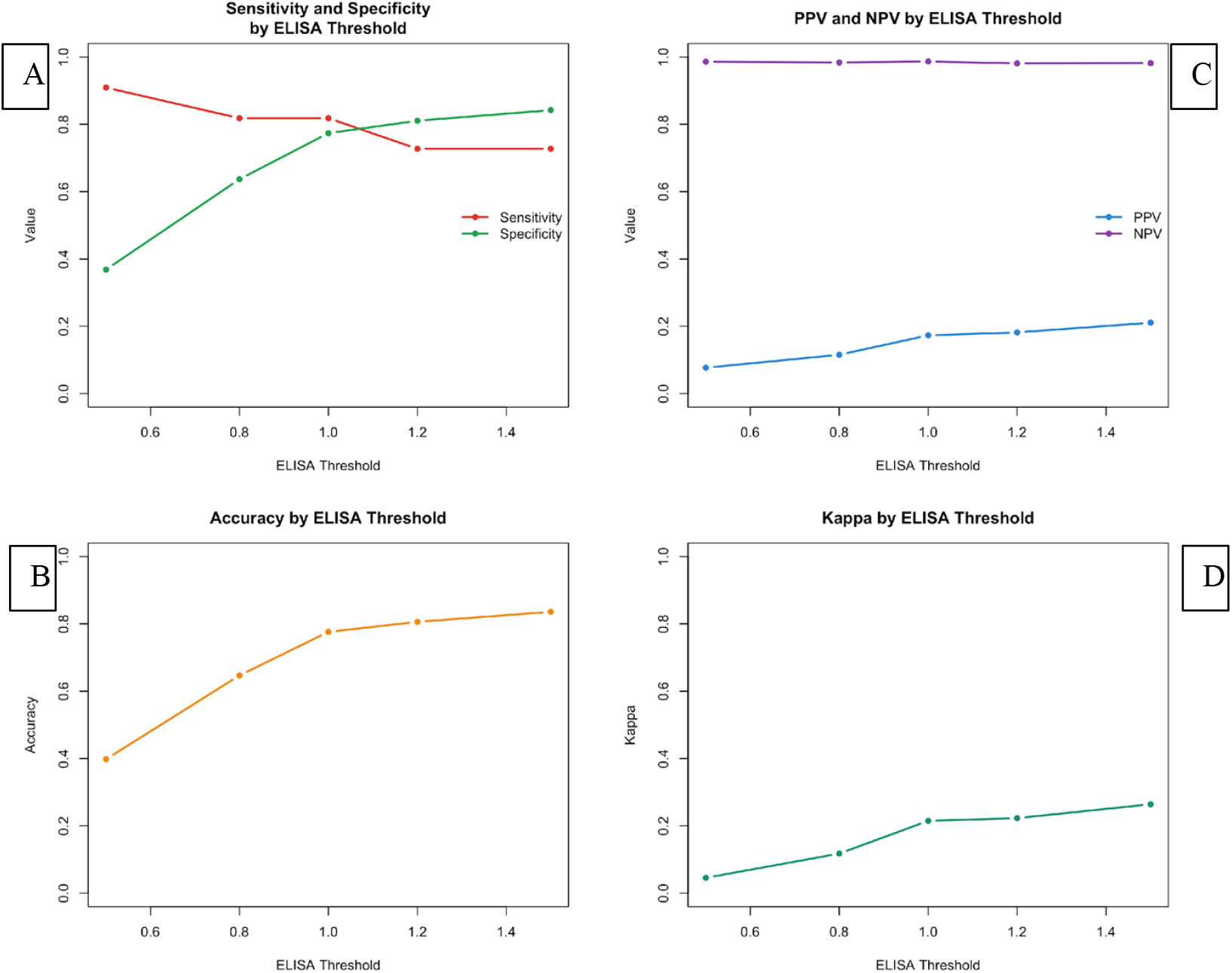
Performance thresholds comparison of ELISA for the detection of Aspergillus antibody.

As expected, sensitivity and specificity demonstrated an inverse relationship across the range of thresholds (Figure 1A). Sensitivity was highest (>95%) at a low threshold (0.2) but at the cost of very low specificity (∼40%). As the threshold increased, sensitivity decreased while specificity improved, reaching near-perfect levels (>99%) at a high threshold (1.4), where sensitivity was negligible.

The overall accuracy (Figure 1B), calculated as the proportion of true results among all tests, was maximized (0.86) at a threshold of 1.0. At this point, the test achieved a sensitivity of 78% and a specificity of 89%.

The Positive Predictive Value (PPV) and Negative Predictive Value (NPV) also varied considerably with the threshold (Figure 1C). The PPV improved steadily with increasing thresholds, exceeding 95% above a threshold of 1.2, indicating that a positive result at this level is highly likely to be a true positive. Conversely, the NPV was highest at low thresholds and remained above 80% until the threshold exceeded 1.0.

The Kappa statistic, which measures agreement beyond chance, reached its maximum value of 0.67 (indicating substantial agreement) at the threshold of 1.0 (Figure 1D), corroborating this as the optimal cut-off for the test under these conditions.

The figure presents a comprehensive analysis of the ELISA test’s performance metrics across a range of potential diagnostic thresholds (0.2 to 1.4).

- **(A) Sensitivity and Specificity:** Illustrates the inverse relationship between sensitivity (solid line) and specificity (dashed line). Lower thresholds favor high sensitivity, while higher thresholds favor high specificity.
- **(B) Overall Accuracy:** Depicts the proportion of all tests that are true positives or true negatives. The peak accuracy (0.86) is achieved at a threshold of 1.0.
- **(C) Positive and Negative Predictive Values (PPV & NPV):** Shows the PPV (solid line) and NPV (dashed line). PPV increases with higher thresholds, whereas NPV is highest at lower thresholds.
- **(D) Kappa Statistic:** Represents the level of agreement between the ELISA test and the reference standard beyond chance. The maximum Kappa value (0.67), indicating substantial agreement, is also observed at the threshold of 1.0.

The vertical reference line across all panels highlights the optimal threshold of 1.0, which provides the best balance between the evaluated metrics.

## Discussion

This study represents the first investigation to document the seroprevalence of Aspergillus-specific antibodies among a post-TB cohort in Senegal, utilizing both ELISA and RDT methodologies within routine healthcare structures. The findings of this study provide crucial initial evidence on the feasibility of integrating serological screening for CPA in a resource-limited setting and highlight the complex interplay between diagnostic performance and implementation practicality.

The central finding is a notably higher seroprevalence detected by ELISA (11.9%) compared to the RDT (5.5%). This trend, where RDTs demonstrate lower sensitivity than ELISA, is consistent with reports from other African settings [10-12]. Studies from Nigeria and South Africa have similarly observed that while RDTs offer excellent specificity, their sensitivity can be variable, potentially leading to an under-ascertainment of true cases in serosurveys [8;13]. The ELISA assay, with its higher analytical sensitivity, likely provides a more accurate estimate of the true antibody prevalence in this population, identifying roughly twice as many seropositive individuals. However, the strong concordance in negative results between the assays suggests that the RDT retains utility as an effective rule-out tool in a screening algorithm.

The choice between these diagnostic modalities has direct implications for implementation. The RDT offers undeniable advantages for decentralized care: its simplicity, rapid turnaround (<30 minutes), and minimal infrastructure requirements make it a transformative tool for point-of-care testing in primary health facilities [5]. This is critical in a context like Senegal, where patients often face significant barriers to accessing central laboratories. Conversely, the ELISA remains the cornerstone for confirmatory testing and robust epidemiological surveillance due to its superior quantitative capacity and sensitivity [7]. Our threshold analysis, which identified an optimal ELISA cut-off that balances sensitivity (78%) and specificity (89%), further strengthens its role as a reference assay.

The clinical significance of our findings is underscored by the study population; individuals with a history of TB, who are at well-documented high risk for developing CPA due to residual cavitary lung lesions [14]. The presence of symptoms like chronic cough and sputum production in our cohort further increases the index of suspicion for fungal sequelae. While our study measures antibody prevalence and cannot confirm clinical CPA in the absence of radiological correlation, the high seropositivity rate strongly suggests a significant burden of Aspergillus sensitization or early disease. This aligns with growing evidence that CPA is a neglected and frequent complication of pulmonary TB in Africa [13; 14].

The interpretation of this seroprevalence findings must be contextualized within the known spectrum of *Aspergillus*-related disease. A positive *Aspergillus* IgG result is a robust indicator of immune sensitization, and in this post-TB cohort, it most strongly suggests a diagnosis of CPA, which aligns with the primary focus of our investigation [3; 15]. However, the antibody response is not exclusive to CPA. It can also manifest in other clinical entities, including *Aspergillus* bronchitis in patients with post-TB bronchiectasis, Allergic Bronchopulmonary Aspergillosis (ABPA), and following other forms of aspergillosis [15; 16]. It is also noteworthy that low-level seropositivity can occur in a minority of asymptomatic individuals, as documented in populations in Uganda and the UK, likely reflecting background environmental exposure [17]; Bongomin et al., 2020 [15]. Therefore, while our data provide critical and novel evidence of a significant *Aspergillus* seroprevalence in Senegal, the definitive attribution to CPA in individual patients requires integration with additional clinical data [3; 18]. Future studies that correlate serological findings with radiological evidence of cavities or fungal balls [19], and with mycological culture or antigen detection [18; 20-23], will be invaluable to precisely define the burden of specific disease entities like CPA within this seropositive population. Furthermore, the incomplete demographic data, particularly from the Yeumbeul site, also limits more detailed subgroup analyses.

Despite these limitations, our findings have a clear programmatic implication: there is an urgent need to integrate CPA screening into post-TB care programs in Senegal and similar settings. A cost-effective, scalable screening algorithm could potentially utilize the RDT for initial rapid screening at peripheral clinics, with positive or equivocal results referred for confirmatory ELISA testing and radiological investigation where available. This would align with WHO recommendations to strengthen fungal diagnostic capacity [24].

In conclusion, this study provides the first sero-epidemiological insight into Aspergillus infection among chronic respiratory patients in Senegal. It confirms the value of antibody testing and underscores the need for a multi-faceted diagnostic approach. Strengthening laboratory capacity for ELISA while simultaneously exploring the deployment of RDTs in a structured screening pathway could significantly improve the early detection and management of this debilitating fungal disease.

## Data Availability

All relevant data are within the manuscript and its Supporting Information files.

## References

1. Denning DW, Cadranel J, Beigelman-Aubry C, Ader F, Chakrabarti A, Blot S, et al. Chronic pulmonary aspergillosis: rationale and clinical guidelines for diagnosis and management. Eur Respir J. 2016;47(1):45–68.

2. Latgé JP, Chamilos G. Aspergillus fumigatus and aspergillosis in 2019. Clin Microbiol Rev. 2020;33(1):e00140–18.

3. González-García P, Alonso-Sardón M, Rodríguez-Alonso B, Almeida H, Romero-Alegría Á, Vega-Rodríguez VJ, et al. How Has the Aspergillosis Case Fatality Rate Changed over the Last Two Decades in Spain? J Fungi. 2022;8(6):576.

4. Hammond EE, McDonald CS, Vestbo J, Denning DW. The global impact of Aspergillus infection on COPD. BMC Pulm Med. 2020;20(1):241.

5. Denning DW. Global incidence and mortality of severe fungal disease. Lancet Infect Dis. 2024;24(7):e428–e438.

6. Sengupta A, Ray A, Upadhyay AD, Izumikawa K, Tashiro M, Kimura Y, et al. Mortality in chronic pulmonary aspergillosis: a systematic review and individual patient data meta-analysis. Lancet Infect Dis. 2025;25(3):312–324.

7. Page ID, Richardson M, Denning DW. Comparison of six Aspergillus-specific IgG assays for the diagnosis of chronic pulmonary aspergillosis (CPA). J Infect. 2016;72(2):240–9.

8. Hunter ES, Page ID, Richardson MD, Denning DW. Evaluation of a new lateral flow device for the serodiagnosis of chronic pulmonary aspergillosis. PLoS One. 2019;14(1):e0210119.

9. Ocansey BK, Otoo B, Gbadamosi H, Afriyie-Mensah JS, Opintan JA, Kosmidis C, et al. Importance of Aspergillus-Specific Antibody Screening for Diagnosis of Chronic Pulmonary Aspergillosis after Tuberculosis Treatment: A Prospective Follow-Up Study in Ghana. J Fungi. 2022;9(1):26.

10. Rozaliyani A, Setianingrum F, Azahra S, Abdullah A, Fatril AE, Rosianawati H, et al. Performance of LDBio Aspergillus WB and ICT Antibody Detection in Chronic Pulmonary Aspergillosis. J Fungi. 2021;7(4):311.

11. Sehgal IS, Soundappan K, Muthu V, Dhooria S, Prasad KT, Rudramurthy SM, et al. Performance of LDBio Aspergillus ICT IgM/IgG Lateral Flow Assay in Diagnosing Chronic Pulmonary Aspergillosis in Community Versus Hospital Setting. Mycopathologia. 2025;190(1):9.

12. Kwizera R, Katende A, Bongomin F, Nakiyingi L, Kirenga BJ. Misdiagnosis of chronic pulmonary aspergillosis as pulmonary tuberculosis at a tertiary care center in Uganda: a case series. J Med Case Rep. 2021;15(1):140.

13. Oladele RO, Otu AA, Richardson MD, Denning DW. Evaluation of a novel handheld volatile organic compound (VOC) monitor for the detection of invasive pulmonary aspergillosis in a UK tertiary hospital. Mycoses. 2018;61(7):469–75.

14. Houbraken J, Kocsubé S, Visagie CM, Yilmaz N, Wang XC, Meijer M, et al. Classification of Aspergillus, Penicillium, Talaromyces and related genera (Eurotiales): An overview of families, genera, subgenera, sections, series and species. Stud Mycol. 2020;95:5–169.

15. Bongomin F. Post-tuberculosis chronic pulmonary aspergillosis: An emerging public health concern. PLoS Pathog. 2020;16(8):e1008742.

16. Barac A, Vujovic A, Drazic A, Stevanovic G, Paglietti B, Lukic K, et al. Diagnosis of Chronic Pulmonary Aspergillosis: Clinical, Radiological or Laboratory? J Fungi. 2023;9(11):1084.

17. Davies AA, Amoah KB, Adjei O, Denning DW, Ayanful-Torgby R. Prevalence of chronic pulmonary aspergillosis in two cohorts of tuberculosis patients in Ghana. Open Forum Infect Dis. 2024;11(4):ofae090.

18. Barac A, Vujovic A, Drazic A, Stevanovic G, Paglietti B, Lukic K, et al. Diagnosis of Chronic Pulmonary Aspergillosis: Clinical, Radiological or Laboratory? J Fungi. 2023;9(11):1084.

19. Nguyen NTB, Le Ngoc H, Nguyen NV, Dinh LV, Nguyen HV, Nguyen HT, et al. Chronic Pulmonary Aspergillosis Situation among Post Tuberculosis Patients in Vietnam: An Observational Study. J Fungi. 2021;7(7):532.

20. Kosmidis C, Denning DW. The clinical spectrum of pulmonary aspergillosis. Thorax. 2015;70(3):270–7.

21. Alvarez-Lerma F, Grau S, Gómez-Álvarez J, Machado M, Nuvials X, Martín-Loeches I, et al. Biomarkers for invasive and chronic aspergillosis: update and future perspectives. Front Cell Infect Microbiol. 2025;15:1599425.

22. Sharma S, Kumar A, Singh S, Goyal A, Khanna M, Ray A. Galactomannan and β-Dglucan as adjuncts to serology in chronic pulmonary aspergillosis. Clin Microbiol Infect. 2024;30(1):64–72.

23. Takazono T, Izumikawa K. Recent Advances in Diagnosing Chronic Pulmonary Aspergillosis. Front Microbiol. 2018;9:1810.

24. World Health Organization. WHO fungal priority pathogens list to guide research, development and public health action. Geneva: World Health Organization; 2022.

